# Decoding Depression Severity from Intracranial Neural Activity

**DOI:** 10.1101/2022.05.19.22275231

**Authors:** Jiayang Xiao, Nicole R. Provenza, Joseph Asfouri, John Myers, Raissa K. Mathura, Brian Metzger, Joshua A. Adkinson, Anusha B. Allawala, Victoria Pirtle, Denise Oswalt, Ben Shofty, Meghan E. Robinson, Sanjay J. Mathew, Wayne K. Goodman, Nader Pouratian, Paul R. Schrater, Ankit B. Patel, Andreas S. Tolias, Kelly R. Bijanki, Xaq Pitkow, Sameer A. Sheth

**Author notes:** Corresponding author: Sameer A. Sheth.

## Abstract

Disorders of mood and cognition are prevalent, disabling, and notoriously difficult to treat. Fueling this challenge in treatment is a significant gap in our understanding of their neurophysiological basis. Here, we used intracranial neural recordings in three patients with severe depression to investigate the neural substrates of this disorder. Across prefrontal regions, we found that reduced depression severity is associated with decreased low-frequency neural activity and increased high-frequency activity. When constraining our model to decode using a single region, spectral changes in the anterior cingulate cortex best predicted depression severity in all three subjects. Relaxing this constraint revealed unique, individual-specific sets of spatio-spectral features predictive of symptom severity, reflecting the heterogeneous nature of depression. The ability to decode depression severity from neural activity increases our fundamental understanding of how depression manifests in the human brain and provides a target neural signature for personalized neuromodulation therapies.

## Main text

Psychiatric and cognitive disorders are among the most challenging ailments we face in terms of social, economic, and public health toll. This challenge derives in large part from their heterogeneity and complexity – heterogeneity in terms of the wide variance in manifestation of these disorders across individuals, complexity in terms of the dearth of objective biomarkers and limited understanding of underlying neurophysiological mechanisms. Adding to their complexity is the fact that these disorders arise from dysfunction not of isolated brain locations but rather of distributed, interconnected networks that span wide-ranging cortical and subcortical regions^1,2^. Networks implicated in psychiatric and cognitive disorders often include prefrontal regions^3–5^, which are the most evolutionarily evolved and are particularly challenging to model in non-human animals. To successfully engage and therapeutically modulate these dysfunctional circuits, we must attain a comprehensive understanding of their pathophysiology. The most precise tools available to accomplish this “circuit dissection” task in humans are electrophysiological recordings and stimulation with intracranial electrodes, given the relatively lower resolution and specificity of non-invasive modalities. Here we apply this approach to investigate the neurophysiological basis of a common and highly burdensome disorder—depression^6^.

Major depressive disorder (MDD) has a lifetime incidence of 10% to 15% and has immense social and economic consequences^7,8^. It is a major contributor to the overall global burden of disease^9^ and in the U.S. alone accounts for more than $200 billion in health care costs^10^. While many conventional treatments are available for depression, nearly one-third of patients are treatment-resistant^11^. A significant number of depressed patients do not respond to first-line medications even after multiple treatment trials^12^. Electroconvulsive therapy and transcranial magnetic stimulation are evidence-based treatments with short-term efficacy, but high rates of relapse are typical^13–15^.

One critical knowledge gap fueling the challenge of treating treatment-resistant depression (TRD) is an insufficient understanding of its neurophysiological basis. Most work to date has used non-invasive methods such as electroencephalography (EEG) and functional MRI^16–20^. These studies have described various alterations in brain activity patterns and suggested potential biomarkers, but precise neurophysiological understanding is still lacking^21^. This understanding, drilled down to the level of the individual, will be essential for treatment-resistant patients being considered for invasive neuromodulation such as deep brain stimulation (DBS)^6,22,23^.

Intracranial recordings such as those performed routinely for seizure monitoring^24^ provide the requisite degree of spatial, temporal, and spectral specificity for this purpose. Whereas studying the neurophysiological basis of mood regulation is possible in the convenience sample of epilepsy patients^25,26^, doing so in a cohort of patients with severe depression and without co-morbid epilepsy would be advantageous. The location of recording electrodes can be targeted to depression-relevant regions, instead of being determined purely for seizure monitoring purposes. Activities during inpatient monitoring can prioritize dense sampling of depression severity measures without concern for interfering with seizure monitoring. The resulting features of interest from the ensuing analyses are more relevant to patients with TRD without contamination by processes related to epilepsy.

Here, our goal is to understand how depression is encoded in the brain by employing an intracranial EEG investigation platform incorporating dense behavioral assessments in TRD patients^6^. In doing so, we seek to address two critical questions: 1) what neurophysiological features characterize depression? 2) can we use these features to reliably predict depression severity? The first question entails finding neural correlates of symptom severity, while the second question addresses the more stringent requirement of finding truly predictive features. As our neuromodulatory therapies advance in sophistication, they will be able to incorporate these biomarkers of health and distress. Doing so will hopefully allow therapeutic supply to more faithfully (spatially and temporally) match symptomatic demand and thereby improve outcomes.

## Results

### Spectral activity across prefrontal regions correlates with depression severity

In our trial, three patients with severe TRD who met eligibility criteria and provided informed consent were implanted with two sets of intracranial electrodes during an initial surgery, one set primarily for stimulating and the other primarily for recording^6,27^ (Fig. 1). The stimulation set consisted of two pairs of permanent DBS leads implanted bilaterally in two regions well-studied in DBS for TRD: the subcallosal cingulate (SCC)^28^ and the ventral capsule/ventral striatum (VC/VS)^29^. The recording set consisted of percutaneously placed temporary “stereo-EEG” ^24^ electrodes implanted in brain regions involved in the regulation of mood and cognition: anterior cingulate cortex (ACC)^18,30,31^, dorsolateral prefrontal cortex (dlPFC)^32,33^, orbitofrontal cortex (OFC)^34–36^, and ventromedial prefrontal cortex (vmPFC)^37,38^ (Fig. 1). Our clinical trial adapted the inpatient intracranial EEG investigation platform commonly used in epilepsy patients, recoined as the neurophysiology monitoring unit (NMU)^6^. Following the initial implant surgery, patients were kept in this inpatient monitoring unit for nine days. We assayed changes in depression severity while simultaneously recording from the implanted electrodes. Clinical outcomes from the first subject in this trial have recently been reported^23^. Here we report neural modeling results and include two additional subjects.

**Fig. 1:**
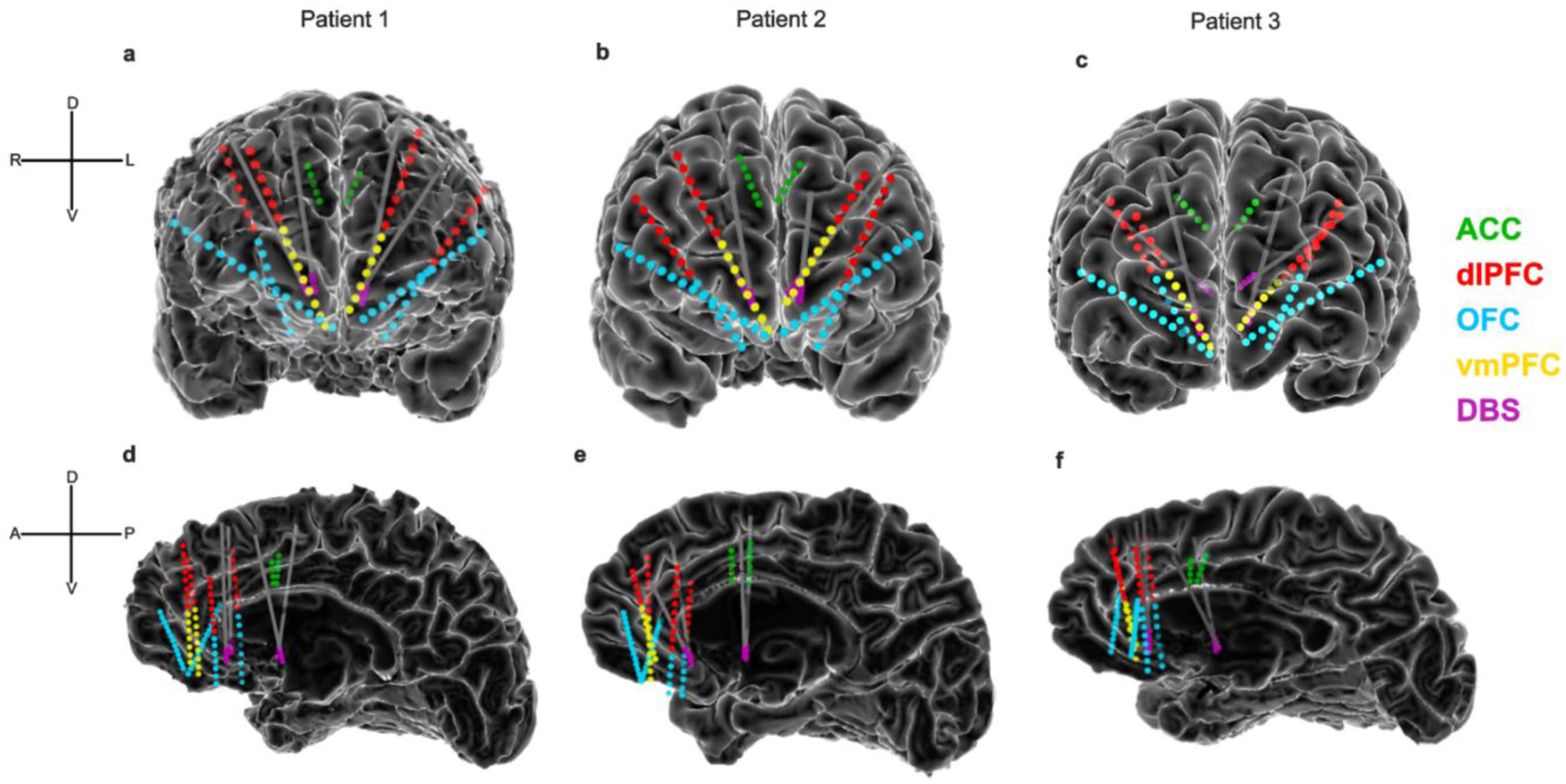
Intracranial recording electrodes sample depression-relevant prefrontal regions. **(a-c)** Frontal views of the reconstructed cortical surface, DBS leads, and stereo-EEG recording contacts for Patient 1, Patient 2 and Patient 3, respectively. In each patient, we implanted stereo-EEG electrodes with the goal of maximizing coverage of key regions while minimizing the total number of electrodes. Stereo-EEG contacts are colored according to the gray matter region sampled: green, anterior cingulate cortex (ACC); red, dorsolateral prefrontal cortex (dlPFC); blue, orbitofrontal cortex (OFC); yellow, ventromedial prefrontal cortex (vmPFC). The contacts of the stimulating DBS leads are shown in purple. (**d-f**) Medial view of the reconstructed cortical surface of the right hemisphere (left hemisphere is hidden for visualization purposes). Dorsal (D), ventral (V), left (L), right (R), anterior (A), and posterior (P) directions are indicated.

We measured symptom severity using the computerized adaptive test depression inventory (CAT-DI), a rapid assessment that correlates with standard depression severity scales^39^. Its adaptive nature allows each administration of this measure to take only 1-2 minutes to complete, and its use of a variety of prompts prevents habituation and provides greater confidence with frequent sampling. We observed substantial variation in depression severity in all three participants (Patient 1: mean severity score= 78.9, std = 6.6; Patient 2: mean severity score = 63.0, std = 4.6; Patient 3: mean severity score = 64.5, std = 14.1) over the 9-day monitoring period, with a trend of declining severity over the NMU stay (Fig. 2a-c).

**Fig. 2:**
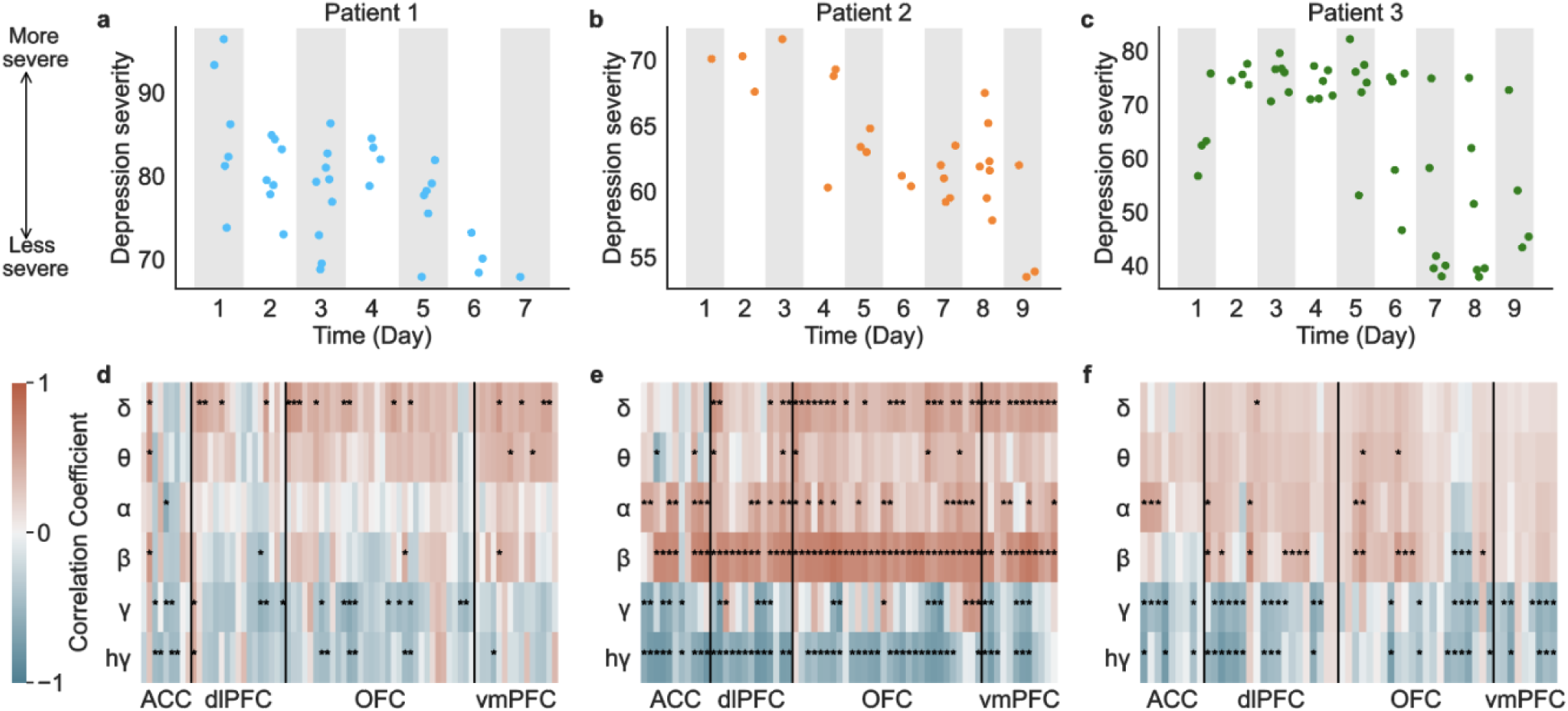
Variation in depression severity captured by neural recordings. (**a-c**) Depression severity (measured by the CAT-DI tool) varied in all three participants. The decreasing trend over time may be related to beneficial effects of stimulation (commonly seen over this time scale), non-specific therapeutic effects of interactions with the research team, and/or other effects. Regardless of the cause(s) of variation, this approach allowed us to frequently sample depression severity over a wide dynamic range while acquiring concurrent dense prefrontal recordings. (**d-f**) Correlation between severity score and brain activity in all gray matter recordings sites. Each horizontal axis increment is a single recording site; the vertical axis shows the frequency bands. The color indicates the correlation coefficient value between severity score and spectral power. A positive correlation (red) indicates that the severity score increases (more severe depression symptoms) when power increases. A negative correlation (blue) indicates that the severity score decreases (less severe symptoms) when power increases. Features with *p* < 0.05 after FDR multiple-testing correction are marked with an asterisk. Although there was inter-individual variability across specific spatio-spectral features, this correlation analysis highlights the trend between low vs. high frequency power and depression severity.

To evaluate how neural activity varied throughout the NMU stay, we extracted spectral power from six frequency bands: delta (1-4 Hz), theta (4–8 Hz), alpha (8–12 Hz), beta (12–30 Hz), gamma (35–50 Hz), and high-gamma (70–150 Hz), yielding six spectral power features per channel for each depression severity measurement. Spectral power features in prefrontal channels showed strong correlations with symptom severity scores after correcting for multiple comparisons. Although there was heterogeneity across patients, power in low frequencies including delta, theta, alpha, and beta were generally positively correlated with symptom severity (Fig. 2d-f, 92.0% of significant features in Patient 1, 99.2% of significant features in Patient 2, and 88.5% of significant features in Patient 3), while power in high frequencies including gamma and high-gamma were generally negatively correlated with symptom severity (92.9% of significant features in Patient 1, 93.2% of significant features in Patient 2, and 100.0% of significant features in Patient 3). In all participants, a majority of brain regions demonstrated decreased low-frequency power and increased high-frequency power when symptoms were less severe.

### Prefrontal neural activity predicts depression severity

The strength of the observed correlations in prefrontal cortex suggests that depression severity may reliably be predicted from spectral power features. To test this hypothesis, we fit regularized regression models to depression severity scores using neural activity recorded across prefrontal sites. Even with the relatively frequent sampling of depression severity, measurements were sparse (Patient 1: 36 measurements; Patient 2: 27 measurements; Patient 3: 47 measurements) relative to the high dimensionality of neural features (approximately 400 features in each patient). In order to increase the generalizability of the model, we reduced the dimensionality of the neural data using automatic region selection and regularized regression. In particular, the model was constrained to use spectral power from a single region to predict depression severity. The selected region was chosen based on the analysis of only training data using a leave-one-out cross-validation strategy. After fitting the model, we used it to predict the depression severity score from a held-out test set and evaluated the prediction error using normalized root mean square error (NRMSE)^26^.

Using this approach, we were able to decode depression severity from neural signals in prefrontal cortex. Our model selected ACC across most folds of cross-validation (all folds in Patient 1 and Patient 2, and 41 out of 47 folds in Patient 3), indicating that ACC was the most informative region for predicting depression severity in all three participants (Fig. 3a-c). Next, we explored relative feature importance within the ACC. Since the predictors were standardized in the model, the regression coefficients for each feature were indicators of feature importance, with larger coefficient magnitudes reflecting greater importance. A few features were consistently more important than others across all folds of the cross-validation and were significantly correlated with symptom severity score (Supplementary Fig. 1). Additionally, there was a strong and significant correlation between predicted and measured symptom score in each participant (Fig. 3d-f; Patient 1: r = 0.66, *p* < 10^−4^, Patient 2: r = 0.93, *p* < 10^−4^, Patient 3: r = 0.68, *p* < 10^−4^), indicating high predictive performance. The decoder was also highly predictive when scores were standardized and pooled across all participants (Fig. 3g; r = 0.73, *p* < 10^−4^).

**Fig. 3:**
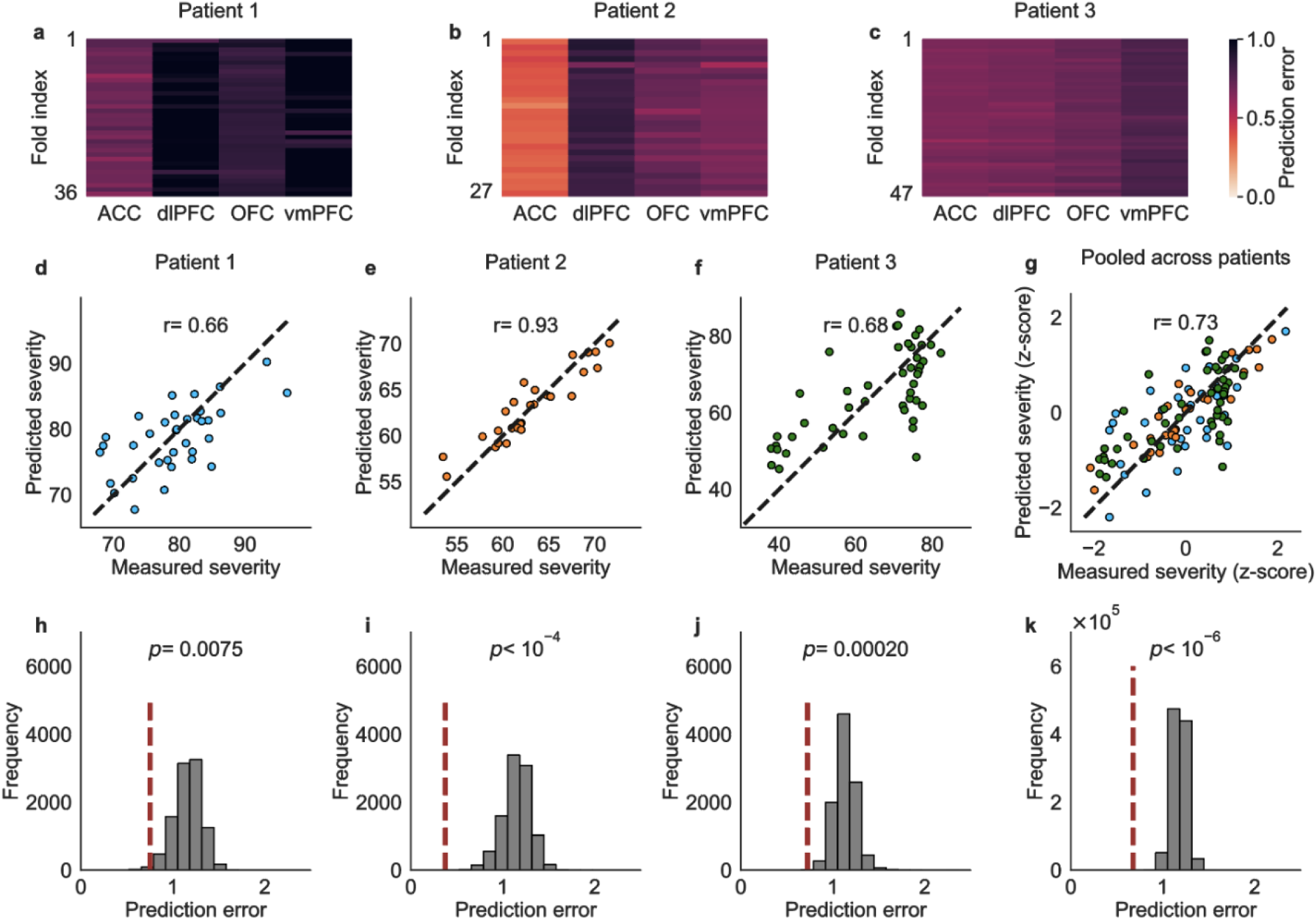
Decoding depression severity from neural activity in prefrontal cortex. (**a-c**) Prediction error of the training set in each fold of the leave-one-out cross-validation. The horizontal axis shows the brain regions, and the vertical axis shows the fold indices of leave-one-out cross-validation. The color indicates the prediction error. Each row includes the prediction error for all regions tested in the training set. Brighter red indicates a smaller error and therefore more accurate prediction. The ACC demonstrated greatest prediction accuracy across most folds of cross-validation, although its degree of supremacy varied across the three subjects. (**d-g**) The predicted score from leave-one-out cross-validation is plotted against the measured score for each depression severity measurement. Points closer to the diagonal indicate more accurate predictions. r values denote Pearson correlation coefficients. Scores from Patient 1 are shown in blue, scores from Patient 2 are shown in orange, and scores from Patient 3 are shown in green. (**h-k**) Distribution of the chance level NRMSE computed from leave-one-out cross-validation for sets of permuted scores (gray, n = 10^4^ permuted tests for each patient and n = 10^6^ for standardized severity scores pooled across patients). NRMSE for the model trained with true measured severity scores is shown as a red vertical dashed line. Permutation testing shows that the true prediction error is significantly lower than chance.

We assessed the significance of the NRMSE with a permutation test to evaluate decoder performance relative to chance. In each patient, we randomly permuted the time indices of severity scores and repeated the same leave-one-out cross-validation process. With the scores permuted and thus mismatched with the neural data, depression severity could no longer be accurately predicted (Supplementary Fig. 2). The prediction accuracies of the decoders were significantly greater than chance performance in all participants (Fig. 3h-j; Patient 1: NRMSE = 0.75, *p* < 0.01, Patient 2: NRMSE = 0.37, *p* < 10^−4^, Patient 3: NRMSE = 0.72, *p* < 10^−3^), and when the scores were pooled across participants (Fig 3k; NRMSE = 0.68, *p* < 10^−6^).

Given our observation of decreasing depression severity over time (Fig. 2a-c), we tested whether our model was trivially identifying a temporal correlation. To do so, we fit a linear regression model to depression severity over time and computed the residuals (Supplementary Fig. 3a-c). This process effectively decorrelated depression severity with time. We then used the neural data to predict the residuals. The decoders accurately predicted the residuals in all patients (Supplementary Fig. 3d-k; *p* < 0.05 for all patients, *p* < 10^−6^ for the scores across patients), indicating that the neural features, not the progress of time, drove the accurate predictions of depression severity.

To add further confidence to our model, we performed 5-fold cross-validation in addition to our original leave-one-out cross-validation approach. The model was trained and selected on four folds of symptom severity scores, and the scores in the other fold were predicted. The decoder accurately predicted depression severity in 5-fold cross-validation as well (Supplementary Fig. 4; *p* < 0.01 for all patients, *p* < 10^−6^ for the scores across patients), providing further confidence in the results.

To evaluate the performance of the region selection technique employed for dimensionality reduction, we fit the decoder without automatic region selection. These decoders can still predict depression severity at levels significantly better than chance (Supplementary Fig. 5a-h; *p* < 0.05 for all patients, *p* < 10^−6^ for the scores across patients), but have larger prediction errors than decoders with region selection (Supplementary Fig. 5i-l). Thus inclusion of region selection improves the decoding accuracy of our model.

Next, we explored the predictability of spectral power from single brain regions and frequency bands (as opposed to all frequency bands) by using these individual features as inputs to the model. While many spectral power features showed significant correlations with depression severity (Fig. 2d-f), not all of these features may necessarily be predictive. We trained separate models for each region and frequency band combination and then performed multiple comparison corrections. For Patient 1, we found that theta, alpha, and beta power in ACC had significant predictability (Fig. 4a). For Patient 2 and Patient 3, we observed more widespread predictability across several regions and frequency bands (Fig. 4b-c). ACC beta power was the overall most predictive feature in Patient 1, whereas ACC high-gamma was most predictive in Patient 2 and Patient 3.

**Fig. 4:**
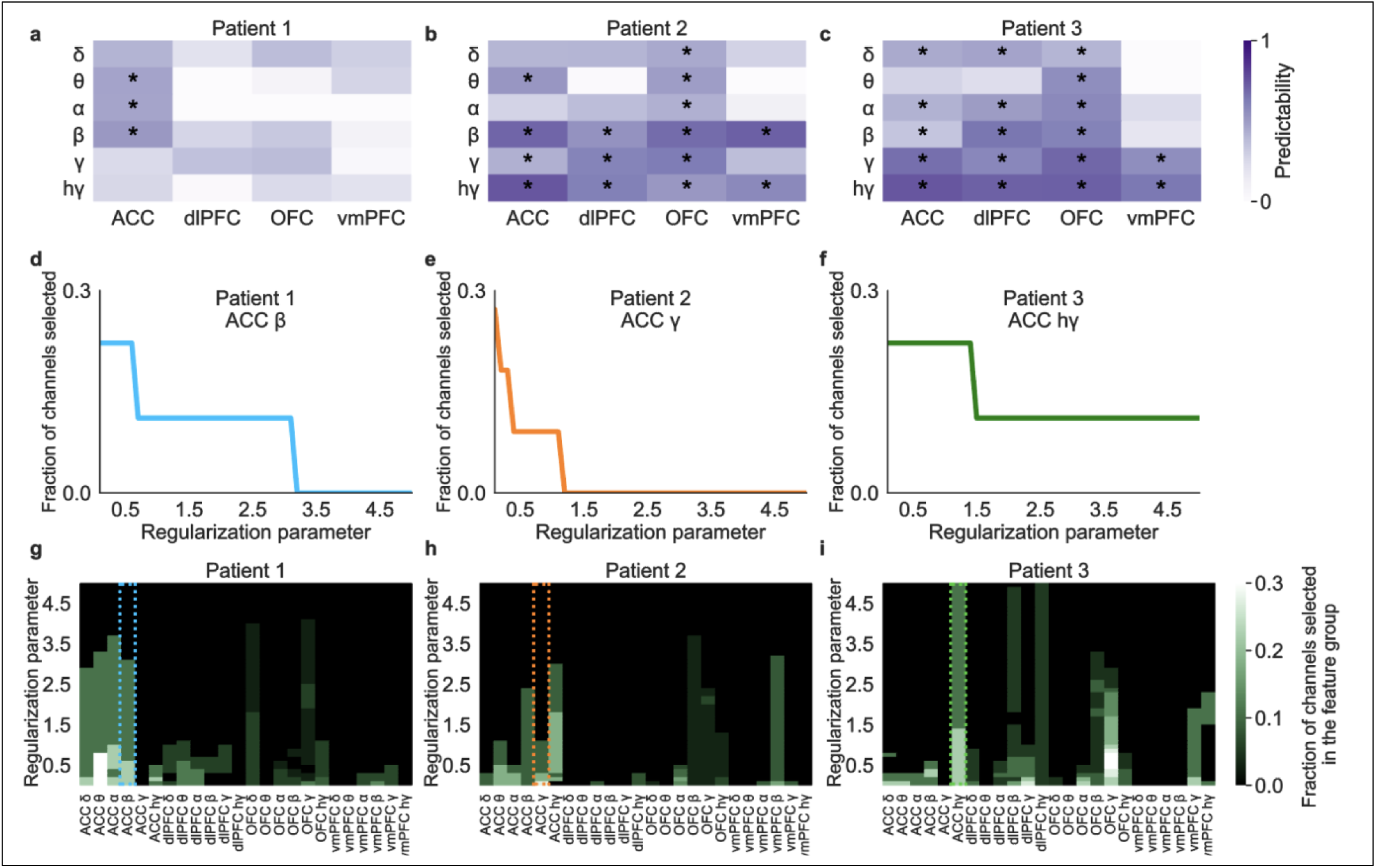
Spatio-spectral features for predicting depression severity are individual-specific. (**a-c**) Predictability of depression severity when using features from a single frequency band in a single brain region. The color indicates the correlation coefficient value between predicted and measured scores. Correlation coefficients with *p* < 0.05 (after FDR correction) are marked with an asterisk. Significantly predictive features are restricted to low-frequency bands in ACC in Patient 1 but are more distributed across spatio-spectral features in Patients 2 and 3. (**d-i**) Fraction of recording channels selected in a specific brain region and frequency band as the regularization parameter alpha varies. (**d-f**) As illustrative examples, we show ACC beta power in Patient 1, ACC gamma power in Patient 2, and ACC high-gamma power in Patient 3. At low values of alpha (more permissive of additional features), many features are selected for inclusion in some fraction of recording channels, but a greater fraction is included for features with most predictive power. As alpha increases, features with most predictive power survive longer and in a greater fraction of channels. (**g-i**) The same information as in **d-f** is shown, but as a heatmap and for all features. The three examples in **d-f** are shown with color-corresponding dotted boxes around their respective column. Again, the fraction of recording channels for particular features decreases with increasing alpha. ACC features are selected the most in these patients, but feature distribution is highly individual-specific.

To further focus on the neural features that are important for predicting depression severity, we fit the model using all spatial and spectral features without automatic region selection. This time, however, we fit the model using a range of values of the regularization parameter alpha to explore the relative importance of individual features. This parameter determines the penalty imposed on the model for including more features. As alpha becomes larger, penalization for adding features increases, and thus fewer features are used. Using ACC beta power, ACC gamma power and ACC high-gamma power as examples, the fraction of recording channels selected in specific feature groups decreases as the regularization parameter alpha increases (Fig. 4d-f). When fitting the model using all individual features, we found that the greatest fraction of channels selected is in ACC, especially when penalization is higher (Fig. 4g-i). As penalization decreases and more features are permitted, spectral features in dlPFC, OFC and vmPFC are increasingly included, indicating that informative features for predicting depression severity are not exclusive to the ACC, but instead are distributed across prefrontal cortical regions in various frequency bands and in an individual-specific manner.

## Discussion

Finding neurophysiological features that characterize and even predict symptom severity is critical for improving our understanding of and developing precise treatments for depression. Here we accurately decoded fluctuations in depression severity over time from intracranially recorded neural activity in three TRD patients. We found that decreased low-frequency power and increased high-frequency power in prefrontal cortex correlate with lower depression severity (i.e., healthier states). Using these spatio-spectral features, we trained a model to predict depression severity and explored the features that were most important for prediction, rather than features that simply showed significant correlations. The transparency and explainability of our model allowed us to identify the ACC as the most influential subregion for decoding depression severity in all three patients. Beyond the ACC, we found various individual-specific neurophysiological features distributed across prefrontal cortex with predictive power. We consider these feature sets to be personalized neural biomarkers for depression severity.

The spectral pattern of correlation we observed, increased high-frequency activity and reduced low-frequency activity, has also been found to be important for decoding positive and negative affect in epilepsy patients^40^. While affective state is not identical to our depression-specific measures, it is likely related. The explainable model we employed was trained on intracranial neural data collected from a cohort of patients with depression, thus eliminating the confounds of epilepsy comorbidities and possible attendant neurophysiological abnormalities. The correlation we observed between neural features and improved depression symptoms is reminiscent of similar associations between this spectral pattern (in particular, increased high-frequency activity) and improved performance in several domains. In visual cortex, for example, increased gamma power predicts faster reaction times in a perception task, perhaps related to increased neural synchrony and resultant facilitated information transfer^41^. Intracranial investigations of human memory have shown that increased high frequency activity and decreased low frequency activity in left temporal cortex predict verbal memory encoding^42–44^, again perhaps reflecting the facilitatory effect of synchronized neuronal spiking^45^. In prefrontal cortex, we have previously shown that increased gamma power predicts improved performance on a cognitive interference task on a trial-by-trial basis^46^, potentially reflecting optimal allocation of cognitive control resources relative to demand^47^. These observations across brain regions mediating perception-to-action behaviors highlight the importance of gamma power as an indicator of neural coherence and a candidate biomarker of performance.

In areas subserving cognitive processes, however, the relationship appears more complicated. Though insufficient application of cognitive resources leads to eroded performance in controlled decision-making tasks^46,47^, unconstrained attentive resources can also be maladaptive. For example, constitutively high activity in PFC regions including the ACC may be the physiological driver of the inappropriately sustained attention to the external environment (e.g., the ordering of objects in a desk drawer) or to internal feelings (e.g., certainty that the stove is off) characteristic of obsessive-compulsive disorder^48,49^. The same requirement for balance is true in affective domains. For example, consider reward sensitivity, one of the cardinal features of positive affect^50^. Whereas a hallmark feature of depression is insufficient reward sensitivity^51^, inappropriately elevated reward-seeking behavior is pathognomonic of addiction disorders^51,52^. Thus in disorders such as depression, which include dysfunction in cognitive and affective domains^53^, these competing forces must achieve balance in order to produce adaptive, productive, euthymic behavior. Consequently, neurophysiological biomarkers of such states will likely be more complex.

We therefore extended our investigation beyond the observed correlations and built a decoder to predict depression severity from spectral power features. Due to the sparse sampling of symptom state relative to that of the electrophysiological data, we began with a model formulation forced to use only the smallest subset of regions, thus producing a model that is more generalizable and less prone to overfitting. The consistent selection of ACC in all patients speaks to the importance of this region in mood and cognitive regulation^40,54^. Even as the region selectivity requirement was relaxed, the prominence of ACC remained (Fig. 4d-i). Although this brain region was commonly selected across patients, the predictive spectral features in ACC differed across participants. The features that were highly predictive were a subset of those that showed significant correlations with depression severity. In Patient 1, beta power was most predictive of depression severity, whereas in Patients 2 and 3, high-gamma power was most predictive. Continued efforts can test the hypothesis that certain features are common predictors across individuals, whereas other features are individual-specific.

As seen in Figure 4, even though the ACC was the most predictive region for a reliable depression decoder in all patients, it was not the only one. Indeed, we found that features with significant predictive power were distributed across various prefrontal regions and frequency bands and were individual specific. This heterogeneity may reflect underlying differences in the involvement of depression-relevant regions across individuals. A growing body of work is attempting to associate differentially involved brain networks with the observed phenotypic diversity of depressive manifestations^2,17^. Within this context, our results further underscore the complex nature of this disorder and the need to account for inter-individual variability in order to optimally engage and treat symptomatic networks^2,17,55,56^.

Future efforts for decoding mood and affect will benefit from the development of continuous, objective behavioral markers. Currently, measuring these domains relies on administering behavioral assessments, a process that suffers from subjectivity and places a high burden on patients to frequently and accurately report their mental state. Contrast this situation with that of motor decoders, which enjoy the advantage of objective and highly temporally resolved measures of position, velocity, acceleration, and related variables^57–60^. Promising methods for affective measures with comparable characteristics include utilizing video and audio recordings^61^. Incorporating facial expression and speech analysis may provide more extensive affective/emotional measures and thereby create the opportunity to develop better mood decoders.

In conclusion, this study demonstrates the feasibility of accurately decoding depression severity based on intracranial prefrontal recordings in TRD patients. With this unique dataset of human intracranial recordings captured alongside measures of symptom severity, we also gain a deeper understanding of the pathophysiology of depression. Ultimately, our findings help to elucidate the neurophysiological underpinnings of depression and may lead to the design of more effective, personalized neuromodulatory interventions.

## Data Availability

All data produced in the present study are available upon reasonable request to the authors

## Acknowledgments

We thank our study participants for their commitment and trust. This study was supported by the National Institutes of Health grant no. UH3 NS103549 (SAS, KRB, JX, NRP, JM, RKM, BM, JAA, ABA, VP, DO, BS, MER, SJM, WKG and NP), grant no. K01 MH116364 (KRB and BM), grant no. R21 NS104953 (KRB and BM), grant no. UH3 NS100549 (WKG), grant no. R01 MH114854 (WKG), and the McNair Foundation (SAS).

## Author contributions statement

SAS and XP initiated the study. JX conceptualized analysis procedures, analyzed the data, and drafted the manuscript with support from NRP, JA, JM, RKM, PRS, ABP, AST, KRB, XP, and SAS. SAS, NP, WKG, SJM, and KRB oversaw the organization of the clinical trial, subject recruitment, and regulatory activities. JX, RKM, BM, JAA, ABA, VP, and DO performed data collection in the neurophysiology monitoring unit. RKM, BS, MER, and KRB performed MRI analysis. XP, SAS, AST, ABP, PRS, and KRB supervised and guided the data analysis. JX, NRP, and SAS wrote the manuscript and all authors contributed to the review and revision of the manuscript.

## Methods

### Study design

An early feasibility trial (NCT03437928) of individualized deep brain stimulation (DBS) guided by intracranial recordings was conducted in adults (n=3) with treatment-resistant depression (TRD). Inclusion criteria include failure of pharmacological, cognitive/behavioral, and electroconvulsive therapies, severity of symptoms, and informed consent. Exclusion criteria include a history of psychosis, personality disorder, recent suicide attempt, or neurodegenerative disorder. All patients (Patient 1, Hispanic male in his 30s; Patient 2, White female in her 50s; Patient 3, White female in her 40s) met inclusion criteria and provided written and verbal consent to participate in the study. This trial is funded by the NIH BRAIN Initiative (UH3 NS103549) and approved by the FDA (IDE number G180300) and our single IRB (Baylor College of Medicine IRB number H-43036).

### Implant surgery

Four DBS leads (Cartesia, Boston Scientific) were placed bilaterally in the ventral capsule/ventral striatum (VC/VS) and subcallosal cingulate (SCC) as previously described^27^. Ten stereoelectroencephalography (sEEG) electrodes were placed bilaterally in depression-relevant brain regions including anterior cingulate cortex (ACC), dorsolateral prefrontal cortex (dlPFC), orbitofrontal cortex (OFC), and ventromedial prefrontal cortex (vmPFC). A stereotactic robotic (ROSA, Zimmer Biomet) was used for the placement of both the DBS leads and sEEG electrodes^6^. Trajectories were carefully planned preoperatively to avoid sulci and blood vessels, and to maximize gray matter coverage. Accurate electrode location was verified intraoperatively using a fluoroscopic computerized tomography (CT) scanner and postoperatively with a true stereotactic CT. Following this implant surgery, patients were kept in the inpatient epilepsy monitoring unit (EMU) while we conducted a series of recording and stimulation studies.

### Depression severity measurements

We acquired measurements of depression severity throughout the nine-day inpatient monitoring period using the computerized adaptive test for depression inventory (CAT-DI)^39^. Our choice of CAT-DI was motivated by our desire to capture symptom states that evolve over relatively short periods of time. Disease states related to mental illness are not static and symptom severity can fluctuate over minutes to days^62^. Depression assessment using standard scales such as the Hamilton Rating Scale for Depression (HAM-D)^63^ and Montgomery-Åsberg Depression Rating Scale (MADRS)^64^ are unwieldy and inappropriate for frequently repeated measurements because they take tens of minutes to administer and are designed for infrequent sampling (days to weeks). The CAT-DI satisfied our need for a dense sampling of depression severity.

Each CAT-DI administration typically includes approximately 12 question items selected from a bank of 389 possible items based on real-time feedback from previous items answered by the participant. Through CAT-DI, we collected rapid and relatively frequent measurements of depression severity. The CAT-DI score has been shown to exhibit a strong correlation with other established depression rating scales such as the Patient Health Questionnaire 9 (PHQ-9) and Hamilton Rating Scale for Depression (HAM-D)^39^. A higher CAT-DI score indicates more severe depression symptoms. A total of 37 CAT-DI tests were completed in Patient 1, 30 CAT-DI tests were completed in Patient 2, and 47 CAT-DI tests were completed in Patient 3 with concomitant neural recordings (see below). For Patient 1, we did not obtain CAT-DI surveys with neural recordings until day 3 of the 9-day period. Therefore, we refer to day 3 after surgery as day 1 in our visualization of CAT-DI scores over time (Fig. 2a).

### MRI and CT imaging protocols

Prior to surgical implant, we conducted a preoperative MRI on a Siemens Prisma 3T scanner with a 64-channel head-neck coil. High-resolution (0.8 mm isotropic) T1-weighted anatomical images (Magnetization-Prepared Rapid Acquisition with Gradient Echo, MPRAGE; repetition time (TR) of 2,400 ms, time echo (TE) of 2.24 ms, an inversion time (TI) of 1,160 ms, a flip angle of 8°, and an acquisition time (TA) of ∼7 min) were acquired. T2-weighted images (SPACE; 0.8 mm isotropic; TR of 3,200 ms, TE of 563 ms, and a TA of ∼6 min) were acquired in the same session. In addition, participants underwent postoperative, high-resolution clinical CT scans to capture electrode placement.

### Cortical reconstructions

FreeSurfer v6.0.0 (https://surfer.nmr.mgh.harvard.edu/)^65^ was used to perform an automatic cortical reconstruction on the preoperative T1-weighted MRI. The T2-weighted MRI was used to improve reconstruction of the pial surfaces. Functional Magnetic Resonance Imaging for the Brain Software Library’s Linear Image Registration Tool (FLIRT) (v6.0)^66,67^ was used to align the postoperative CT data to the preoperative T1-weighted MRI. The postoperative CT data were used to determine the contact positions relative to local neuroanatomy.

### Electrode localization

Electrode coordinates were determined manually from the co-registered CT data in BioImage Suite v3.5b1^68^ and placed into the native MRI space. An expert rater (BS) examined the images and determined whether the contact was in white or gray matter based on where it was plotted on the brain slice. Contacts that were determined to be in white matter were excluded from further analysis. Electrodes were also manually labeled to brain regions according to their anatomical location by the same rater. The cortical surface (pial surface) and electrode locations were reconstructed using the Multi-Modal Visualization Tool^69^.

### Intracranial recordings

Neural signals were recorded during the administration of CAT-DI test. Herein we will refer to each CAT-DI timepoint and its associated neural data as a ‘block’. Signals were recorded with sEEG electrodes at 2 kHz using a Cerebus data acquisition system (BlackRock Microsystems, UT, USA). All signals were amplified and bandpass filtered from 0.3-500 Hz (4th order Butterworth filter). DBS was off during all recordings.

### Neural data preprocessing and signal conditioning

Raw signals were visually inspected for the presence of recording artifacts. Channels that were found to have excessive noise were excluded to prevent noise from spreading to other channels through re-reference. Blocks with poor quality of the neural recording in more than half of the channels were excluded from further analysis. One block out of 37 was removed from Patient 1, three blocks out of 30 were removed from Patient 2, and no block was removed from Patient 3. Each channel was notch filtered (60 Hz and its harmonics) to reduce line-noise artifacts. To reduce the effects of volume conduction, signals were then re-referenced through bipolar reference by subtracting the voltage of the neighboring contact on each sEEG electrode^70^. A total of 75 sEEG gray matter channels in Patient 1, 66 channels in Patient 2, and 59 channels in Patient 3 remained after bipolar referencing were used for further analysis.

### Feature extraction

After signals were down-sampled to 1000 Hz, we performed a Hilbert transform to estimate spectral power features in six different frequency bands: 1-4 Hz (delta), 4–8 Hz (theta), 8–12 Hz (alpha), 12–30 Hz (beta), 35–50 Hz (gamma) and 70–150 Hz (high-gamma). Then for each channel, we log-transformed the average power during the CAT-DI test for each frequency band.

### Spectral activity analysis

Pearson correlation coefficient and corresponding *p*-value were calculated between depression severity score and spectral power features from six frequency bands in all gray matter channels. FDR multiple-testing correction was performed across the frequency bands and channels to correct *p*-values. A Pearson correlation coefficient with corrected *p*-value smaller than 0.05 was defined as significant.

### Automatic region selection

The recording channels per patient were distributed across four main prefrontal regions: ACC, dlPFC, OFC, and vmPFC. To increase the generalizability of the model and avoid overfitting, we greatly reduced the number of model parameters by using a region selection technique^26^. Automatic region selection was employed to minimize the number of regions used in the decoder. Using training data only, we implemented model selection through inner cross-validation. The optimal brain region for decoding was determined by comparing independent models, each fit with the channels in one specific region. In the first stage of candidate model selection, independent decoders were fit with a single region, and one was selected. If the selected decoder achieved significance, automatic region selection stopped. Otherwise, the next stage consisted of fitting models using channels from two regions. The process continued until a decoder was found that achieved significance. In all three patients, significance was achieved after only the first stage, using features from a single region. Thus, this process largely decreased the number of model parameters.

### Model fitting

We used LASSO regression to fit neural data to the CAT-DI scores as described in (1). LASSO regularization was used to minimize the objective function:

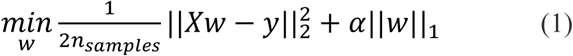

where *y* is the measured CAT-DI score and *X* includes all features in the optimal brain region, which is selected by automatic region selection with inner-level cross-validation. *w* is the weight vector. 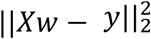 is the residual sum of squares, which measures the discrepancy between the predicted scores and the measured scores. ||*w*||_1_ is the ℓ1 term, which is the sum of the magnitudes of weights in the weight vector. *α* is a constant that multiplies the ℓ1 term and is selected using inner cross-validation. By adding ℓ1 penalty, this regularization encourages sparsity since coefficients for many features are likely to become zero, hence eliminating these features from the model. Coordinate descent was used to fit the weights^71^. This regression method further reduces the dimensionality of the model.

### Measurement of prediction error

We quantified the prediction error using normalized root mean square error (NRMSE)^26^ as described in equation (2). NRMSE is defined by

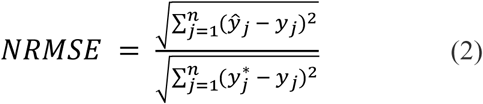

where *n* is the total number of CAT-DI scores, *ŷ*_*j*_ is the *j*th predicted score, *y*_*j*_ is the *j*th measured score, and 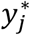 is the mean of other measured scores except for the *j*th measured score. The NRMSE quantifies how much more accurately the model predicts a test score relative to a model that predicts a test score simply as the average of the other scores. A lower NRMSE indicates that the prediction is more accurate. NRMSE was used as the prediction error criterion for selecting candidate models and for the final evaluation of the selected model when testing model significance.

### Cross-validation

To assess the performance of our decoder, we used both leave-one-out (Fig. 3) and 5-fold cross-validation (Supplementary Fig. 4).

For leave-one-out cross-validation, we left out one CAT-DI score as test data and used the rest of the scores as training data to select the region and build the model. Through inner cross-validation using only the training data, we obtained the prediction error for all candidate models. To make the model more robust to noise, we defined a feature value as an outlier when it was more than four standard deviations away from the mean value of that feature in the training set. We then automatically substituted all outlier values with the mean values of the corresponding feature in the training data. Less than 2% of feature values were identified as outliers for all participants.

The model with the lowest prediction error was then selected as the best model. This model fitting and selection process had no knowledge of the test CAT-DI score. We then used the best candidate model from each fold to predict the corresponding test score in the outer cross-validation. We repeated this leave-one-out procedure for all CAT-DI scores and then computed the NRMSE of the decoder using equation (2).

For 5-fold cross-validation, we split the data into five folds, trained the model on four folds of CAT-DI scores, and predicted the scores in the other fold. We repeated this procedure five times to obtain predicted scores for all CAT-DI points and then computed NRMSE to evaluate the performance of the decoder. In both leave-one-out and 5-fold cross-validation, model training and selection were conducted using only CAT-DI scores and neural activity within the training set.

### Model assessment

We first calculated the true prediction error using the measured scores and predicted scores from each patient. In order to estimate decoding performance due to chance, we randomly permuted each symptom severity score on the neural data and then repeated the cross-validation process in the same manner as before. We did 10^4^ random permutations to build a distribution of prediction errors expected due to chance. Next, we counted the number of samples in the distribution for which the prediction error due to chance was lower than the true prediction error. In this way, the *p*-value was defined as the probability that a model fit to a permuted set of symptom severity scores had a higher prediction accuracy than a model fit to the true scores.

Next, we evaluated the model across all patients by pooling their data. CAT-DI scores were z-scored based on the measured scores within each patient. These standardized scores were then pooled across patients. We repeated the permutation process 10^6^ times to estimate the chance distribution of pooled prediction errors and calculated the corresponding *p*-value in the same way as described above. Here, the significance of the prediction error was the probability that decoding using the permuted set of CAT-DI scores (z-scored) has a lower NRMSE than using the true set of CAT-DI scores (z-scored).

### Evaluation of time as a potential confound of model performance

CAT-DI scores for all three participants decreased over days in the EMU (Fig. 2a-c). In order to ensure that neural features were truly capturing fluctuations in depression severity rather than the passage of time, we fit a linear regression model to depression severity over time using the least-squares approach. For each data point, we calculated the residual by subtracting the predicted depression severity score using the line of best fit from the actual severity score. In this way, the resulting residuals were decorrelated with time (Supplementary Fig. 3). We then used the neural activity to predict the residuals using the same leave-one-out cross-validation technique as we have previously described.

### Evaluation of feature importance

In order to evaluate the relative importance of features corresponding to individual frequency bands and subregions, we fit the model using features from single subregions and single frequency bands. We then calculated the Pearson correlation coefficient between predicted and true severity scores, and computed the corresponding *p*-values with FDR multiple-testing correction.

Lastly, to investigate feature importance without automatic region selection, we fit all features with LASSO regression at a range of different regularization parameter values. The regularization parameter alpha determines the penalty for using more features. As alpha decreases, the number of features used in the model increases. We fit the model using 50 different alpha values ranging from 0.1 to 5.0 in steps of 0.1. At each step, we calculated the fraction of channels in each feature group (defined by the region and frequency band) selected by the model. We completed these analyses using all data, as our goal was to better understand the effects of variation in regularization parameter alpha on the features that were selected by the model.

## Supplementary Figures

**Supplementary Fig. 1:**
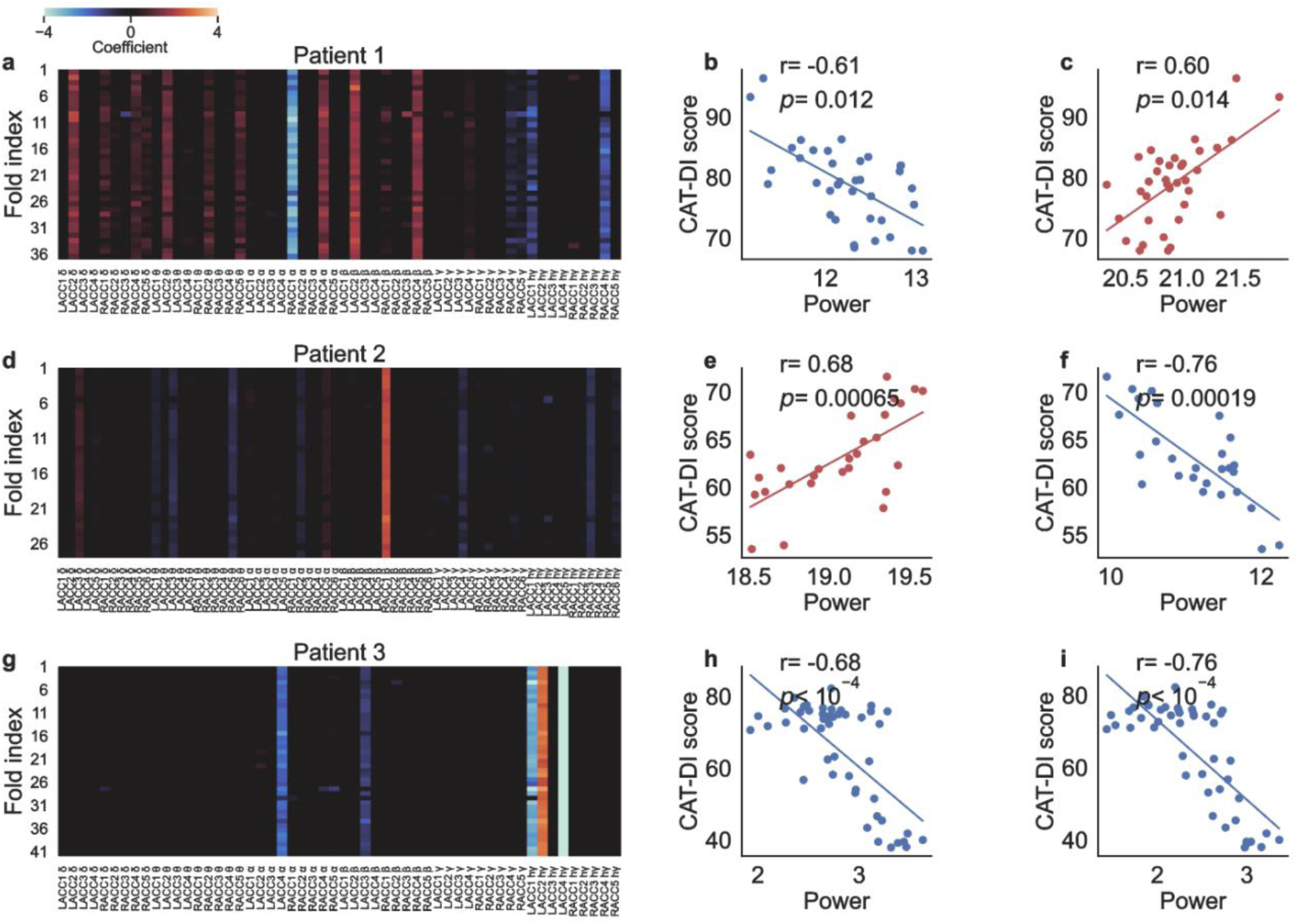
Training data is used to select features that are predictive of depression severity. (**a, d, g**) LASSO regression coefficients for input features in each fold of the leave-one-out cross-validation selecting ACC. Horizontal axis shows the feature names. Vertical axis shows the fold indices of leave-one-out cross-validation. The color indicates the coefficient value. Each row includes the coefficients for all features in the selected region from that fold. Most coefficients become zero in the LASSO regression, thus the color in most areas is black. Red indicates that the coefficient in the LASSO regression is positive, and blue indicates that the coefficient is negative. (**b-c, e-f, h-i**) The correlation coefficient and corresponding *p*-value (after FDR multiple-testing correction) between the CAT-DI score and the value of example features. Red indicates that the correlation coefficient is positive, and blue indicates that the correlation coefficient is negative.

**Supplementary Fig. 2:**
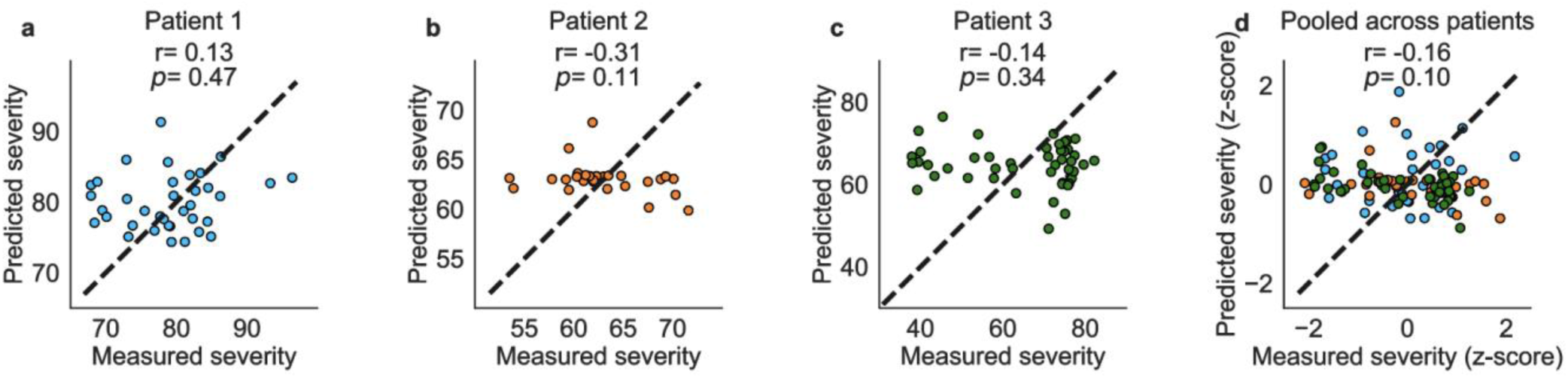
Permuted CAT-DI scores yield low predictive performance. Predictions for an example set of permuted CAT-DI scores in Patient 1 (**a**), Patient 2 (**b**), Patient 3 (**c**), and pooled across patients (**d**).

**Supplementary Fig. 3:**
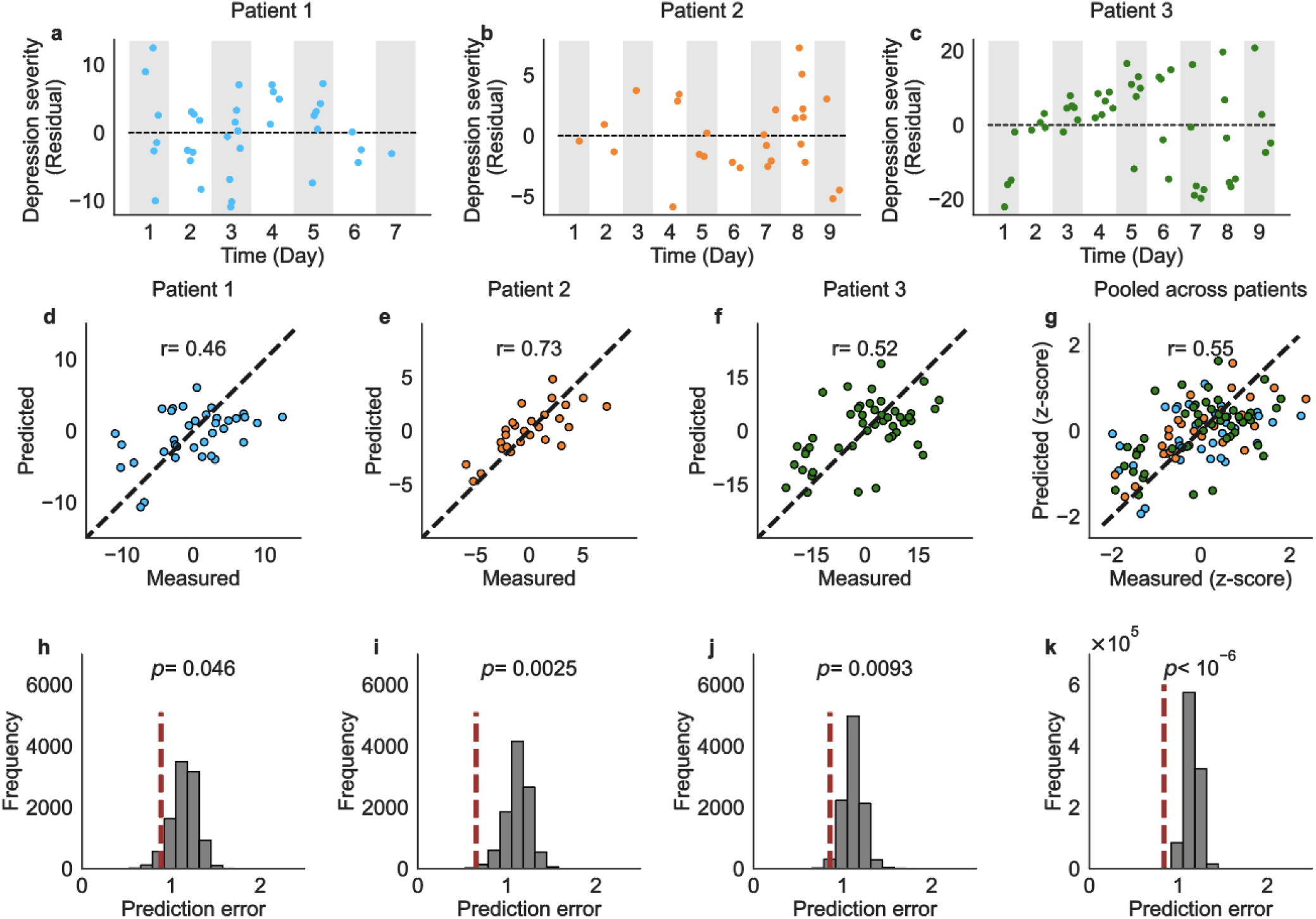
Depression severity can be predicted after removing the effect of time. (**a-c**) We fit a linear regression model between depression severity and time using the least-squares approach. For each data point, we calculated the residual by taking the difference between the actual depression severity and the predicted severity from the line of best fit. The residuals were then predicted by the neural activity. (**d-g**) The predicted residual from leave-one-out cross-validation is plotted against the true residual. (**h-k**) Distribution of the NRMSE for sets of permuted residuals (gray, n = 10^4^ permuted tests for each patient and n = 10^6^ for z-scores pooled across patients). NRMSE for the model trained with true residuals is shown as a red vertical dotted line.

**Supplementary Fig. 4:**
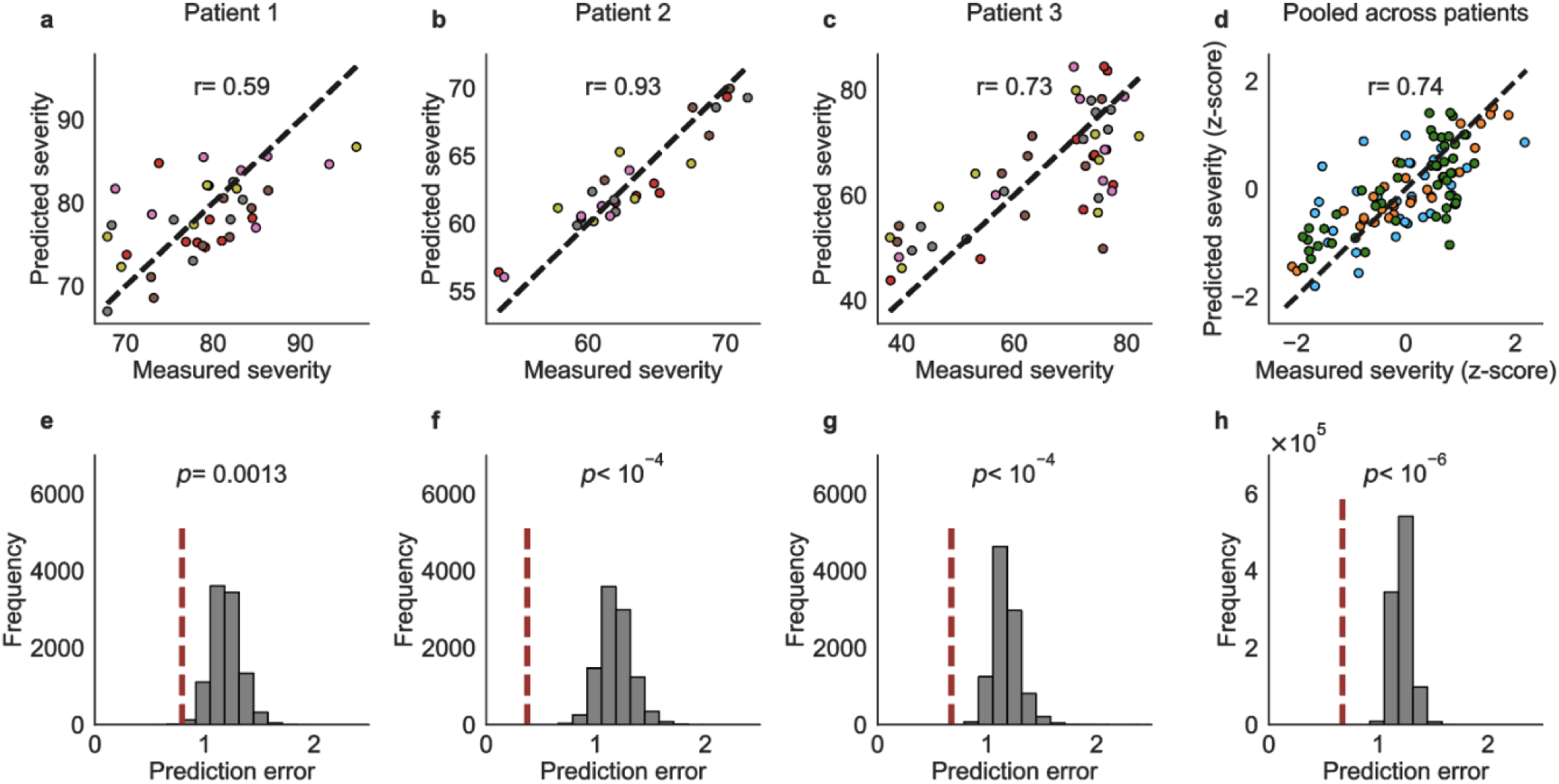
Depression severity can be decoded using 5-fold cross-validation. (**a-d**) The predicted score from each fold of 5-fold cross-validation against the measured test score corresponding to that fold. Points with the same color come from the same fold. Scores from Patient 1 are shown in blue, scores from Patient 2 are shown in orange, scores from Patient 3 are shown in green in **d**. The Pearson correlation coefficients are shown on the plots. The corresponding *p*-value is less than 10^−4^ for each patient and pooled across patients. (**e-h**) Distribution of the NRMSE for sets of permuted scores (gray, n = 10^4^ permuted tests for each patient in 5-fold cross-validation and n = 10^6^ for z-scores pooled across patients). NRMSE of the model trained using true measured CAT-DI scores is shown as a red vertical dotted line. Permutation testing shows a prediction error significantly lower than chance in 5-fold cross-validation in Patient 1, Patient 2, Patient 3, and when the scores were pooled across patients.

**Supplementary Fig. 5:**
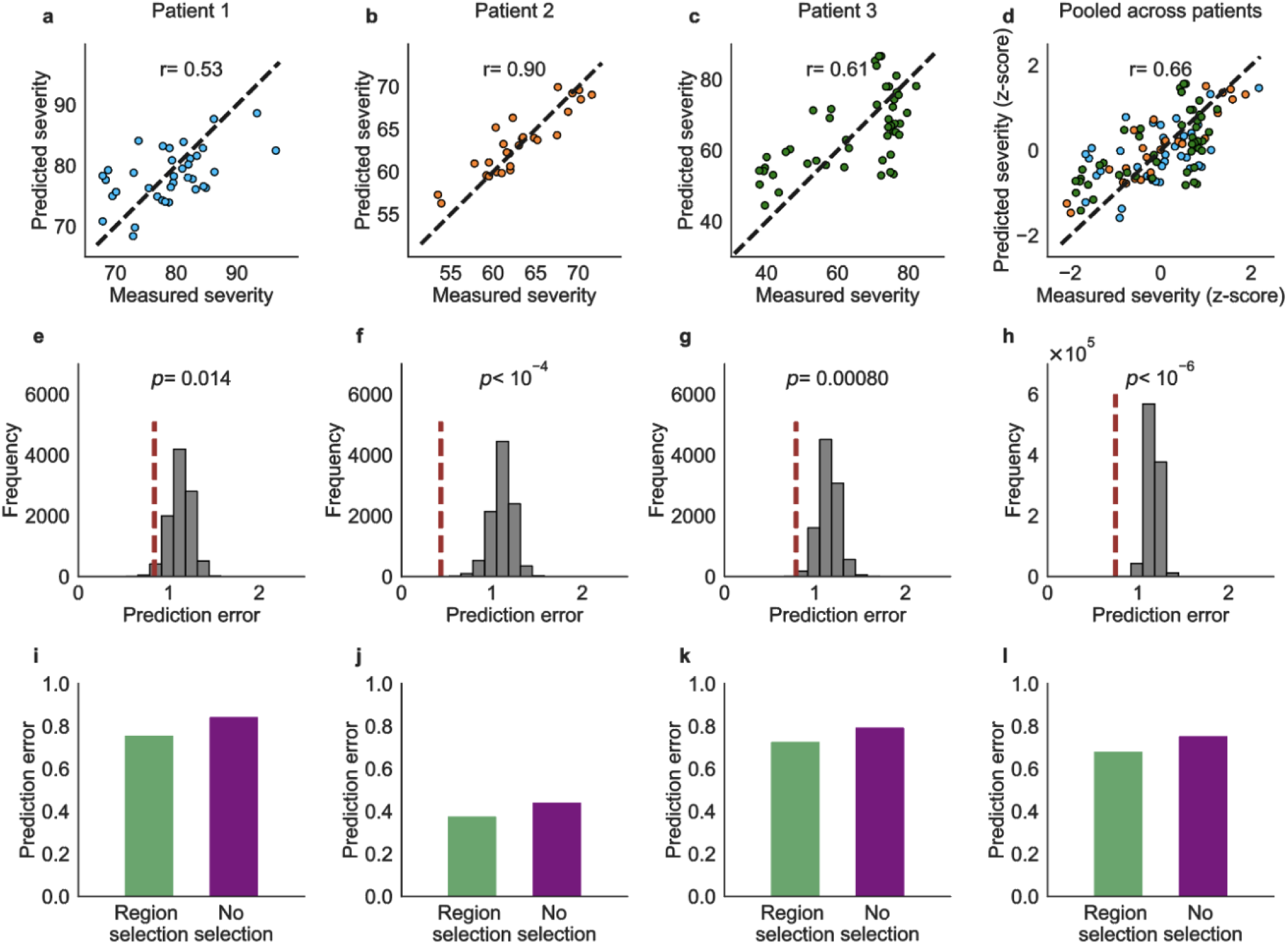
Predictability when decoding without automatic region selection. (**a-d**) The predicted score from leave-one-out cross-validation against the measured score when using LASSO regression without automatic region selection. (**e-h**) NRMSE of the LASSO regression model trained using true measured CAT-DI scores (red vertical dotted line) and distribution of the NRMSE for sets of permuted scores (gray bars, n = 10^4^ permuted tests for each patient, n = 10^6^ permuted tests for z-scores pooled across patients). (**i-l**) Comparison between prediction error in models with and without region selection technique. A lower prediction error indicates a more accurate prediction.

